# Retrospective screening of routine respiratory samples revealed undetected community transmission and missed intervention opportunities for SARS-CoV-2 in the United Kingdom

**DOI:** 10.1101/2020.08.18.20174623

**Authors:** Joseph G. Chappell, Theocharis Tsoleridis, Gemma Clark, Louise Berry, Nadine Holmes, Christopher Moore, Matthew Carlile, Fei Sang, Johnny Debebe, Victoria Wright, William L. Irving, Brian J. Thomson, Timothy C.J. Boswell, Iona Willingham, Amelia Joseph, Wendy Smith, Manjinder Khakh, Vicki M. Fleming, Michelle M. Lister, Hannah C. Howson-Wells, Edward C. Holmes, Matthew W. Loose, Jonathan K. Ball, C. Patrick McClure, on behalf of the COG-UK consortium

## Abstract

In the early phases of the SARS coronavirus type 2 (SARS-CoV-2) pandemic, testing focused on individuals fitting a strict case definition involving a limited set of symptoms together with an identified epidemiological risk, such as contact with an infected individual or travel to a high-risk area. To assess whether this impaired our ability to detect and control early introductions of the virus into the UK, we PCR-tested archival specimens collected on admission to a large UK teaching hospital who retrospectively were identified as having a clinical presentation compatible with COVID-19. In addition, we screened available archival specimens submitted for respiratory virus diagnosis, and dating back to early January 2020, for the presence of SARS-CoV-2 RNA. Our data provides evidence for widespread community circulation of SARS-CoV2 in early February 2020 and into March that was undetected at the time due to restrictive case definitions informing testing policy. Genome sequence data showed that many of these early cases were infected with a distinct lineage of the virus. Sequences obtained from the first officially recorded case in Nottinghamshire - a traveller returning from Daegu, South Korea – also clustered with these early UK sequences suggesting acquisition of the virus occurred in the UK and not Daegu. Analysis of a larger sample of sequences obtained in the Nottinghamshire area revealed multiple viral introductions, mainly in late February and through March. These data highlight the importance of timely and extensive community testing to prevent future widespread transmission of the virus.

## Introduction

Severe acute respiratory syndrome coronavirus 2 (SARS-CoV-2) is a novel zoonotic virus, first identified in the city of Wuhan in the Chinese province of Hubei, following a cluster of patients presenting with severe pneumonia ^1^. Since this first detection in December 2019, SARS-CoV-2 has rapidly spread across the globe and, as of 3^rd^ August 2020, there have a been a total of 17,918,582 confirmed cases globally, resulting in 686,703 deaths ^2^.

Infection with SARS-CoV-2 can lead to the development of coronavirus disease 2019 (COVID-19), characterised by fever, persistent cough, fatigue and shortness of breath ^3,4^. In severe cases, this can progress into acute respiratory distress syndrome (ARDS), often requiring artificial ventilation^5^, although many cases also present asymptomatically or with only mild disease. Asymptomatic and pre-symptomatic carriage of SARS-CoV-2 is now well documented ^6,7^ and transmission has been reported ^8–10^, which is thought to be related to high levels of viral shedding in the upper respiratory tract during the early stages of infection ^11^. Due to the difficulty in identifying infected asymptomatic or pre-symptomatic individuals, SARS-CoV-2 has been able to rapidly spread, particularly in healthcare and age care environments ^12^.

The first confirmed SARS-CoV-2 case in the United Kingdom travelled from Hubei province on the 23^rd^ of January 2020 and became symptomatic on the 26^th^ of January. This patient then transmitted the virus to a household contact, who also became symptomatic two days later ^13^. The third diagnosed case was in a traveller returning from Singapore on the 6^th^ of February; this patient had stopped in France where they infected seven others, before travelling to and seeding several infections in the UK^14^. However, retrospective analysis of genome and associated metadata has conservatively estimated a minimum of 1356 independent introductions of the virus into the UK, primarily from travellers originating in Spain, France or Italy during mid-March^15^. There have been 304,699 lab-confirmed cases and 46,201 deaths, in the UK as of the 3rd of August 2020 ^2^.

Initial SARS-CoV-2 RT-PCR testing in the UK was offered via referral to Public Health England (PHE) national and regional diagnostic laboratories, and required strict epidemiological and clinical criteria to be met, specifically a recent travel history to Hubei province or contact with a known case and 1 or more of fever, shortness of breath or new and persistent dry cough. These case definitions were revised on several occasions to include travel to mainland China and several other Asian countries initially (7^th^ of February), then expanded further to include Iran and northern Italy (25^th^ of February) before finally being removed as essential criteria for diagnostic testing on the 12^th^ of March ^16^. Importantly, a case definition that relied heavily on travel history or exposure to a known infected individual likely resulted in undetected cases and transmission in both healthcare and community settings. A broadening of epidemiological criteria in COVID-19 case definitions was associated with an increased proportion of COVID-19 cases being identified ^17^. Furthermore, as SARS-CoV-2 testing was initially only available via PHE laboratories, and testing within NHS laboratories was not rolled out until March 2020, this further restricted the capacity to detect early cases and transmission events.

To better understand the prevalence and emergence of SARS-CoV-2 in the UK before the broadening of case definition criteria and wider testing, we conducted a retrospective screening of case histories to identify individuals with symptoms compatible with SARS-CoV-2 infection, as well as retrospective PCR testing of archived diagnostic specimens submitted for respiratory virus screening. This study was conducted in a large teaching hospital located in Nottingham and representative of provincial cities throughout the UK. We describe the detection of SARS-CoV-2 from 8 patients admitted to hospital with severe respiratory distress who were not tested at the time because they had no travel history or contact with someone infected and therefore did not meet the case definition applied at the time. Sequence analysis of these early cases showed that they belonged to a distinct B-lineage of SARS-CoV-2 which dominated the early phases of the local outbreak. Analysis of further sequences, collected as part of the COG-UK initiative ^1^ from patients who tested positive after the rollout of local testing, highlighted extensive introductions of the virus into the region.

## Methods

### Sample collection

A total of 1660 respiratory specimens (throat swabs, nose swabs, nasopharyngeal aspirates, bronchoalveolar lavages and endotracheal tube secretions) from 1378 patients were collected between the 2^nd^ of January and 11^th^ of March 2020 for routine diagnostic investigation^19^ for which surplus total nucleic acid was available. To facilitate rapid testing, we created 169 pools each containing up to 10 samples ^19^, which were then subjected to SARS-CoV-2-specific PCR.

### RT-PCR screening and high-throughput sequence analysis

cDNA was synthesised from each of the nucleic acid pools using RNA to cDNA EcoDry (Random Hexamers) (Takara Bio Europe, Saint-Germain-en-Laye, France). The cDNA was initially screened with an in-house PCR assay targeting the RNA-dependent RNA polymerase (RdRp) gene region of SARS-CoV-2, producing an 186bp amplicon. Positive samples were then confirmed using a larger 366 bp amplicon assay, also targeting the RdRp. For both assays, 5 μl of cDNA was added to PCR reactions containing 5 μl of 10x PCR buffer, 0.25 μl of HotStarTaq DNA Polymerase (QIAGEN, Hilden, Germany), 2 μl each of both forward and reverse primers (10 pmol/μl), 2 μl dNTPs (10 mM) (Sigma-Aldrich, St Louis, USA) and 33.75 μl of DEPC-treated water. Cycling conditions were 95 °C for 15 min, 55 cycles of 95°C, 58°C and 72°C for 20s each, followed by a final extension of 72°C for 30 s. The primer sequences for the initial 186bp assay was qCOV19f: 5’- CAATAGCCGCCACTAGAGGA & qCOV19r: 5’- GAGCAAGAACAAGTGAGGCC. The sequences for the larger assays were COV19_Cf: 5’- CGCCACTAGAGGAGCTACTG and COV19_Cr: 5’-GCCGTGACAGCTTGACAAAT.

Positive pools were de-multiplexed and the individual nucleic acid extracts were subject to the same methodology as the pooled samples. Whole-genome sequencing was achieved using the ARTIC amplicon sequencing protocol ^20^, the complete methodology is available in the supplementary information.

### Phylogenetic Analysis

In total, 28,124 SARS-CoV-2 whole genomes with >95% coverage were downloaded from GISAID on June 10^th^, 2020. The processing of the genomes was performed with the Geneious Prime 2019.0.4 software. The genomes were sub-divided based on their lineage as indicated by the Pangolin tool ^21^. For each lineage, the sequences were aligned and the 5’ and 3’ ends were trimmed. Maximum likelihood phylogenetic trees were generated for multiple lineages (B.1, B.1.1, B.1.1.1, B.1.p11, B.1.5, B.1.34, B.1.36, B.2, B.2.1, B.2.2, B.2.5, B.2.6, B.3, B.6, B.9, B.10, B.15) using IQ-TREE2 ^22^ employing the GTR+F+R3 model of nucleotide substitution as suggested by the software’s model finder, with 1000 SH-like approximate likelihood ratio test (SH-aLRT)^23^. Each tree was rooted using the early Wuhan virus hCoV-19/Wuhan/WH04/20201 EPI_ISL_40680112020-01-05 that was sampled in January 2020.

## Results

### Clinical Diagnosis of SARS-CoV-2

Prior to the rollout of localised SARS-CoV-2 testing on the 12^th^ of March 2020, 9 cases (denoted Patients 9-17) matching the clinical and epidemiological contemporary case definitions were identified and referred to PHE for testing (Supplementary Table 1). Complete sequence data was available for one patient (Patient 9), a traveller returning from South Korea, tested on the 28th of February (Table 1).

**Table 1.**
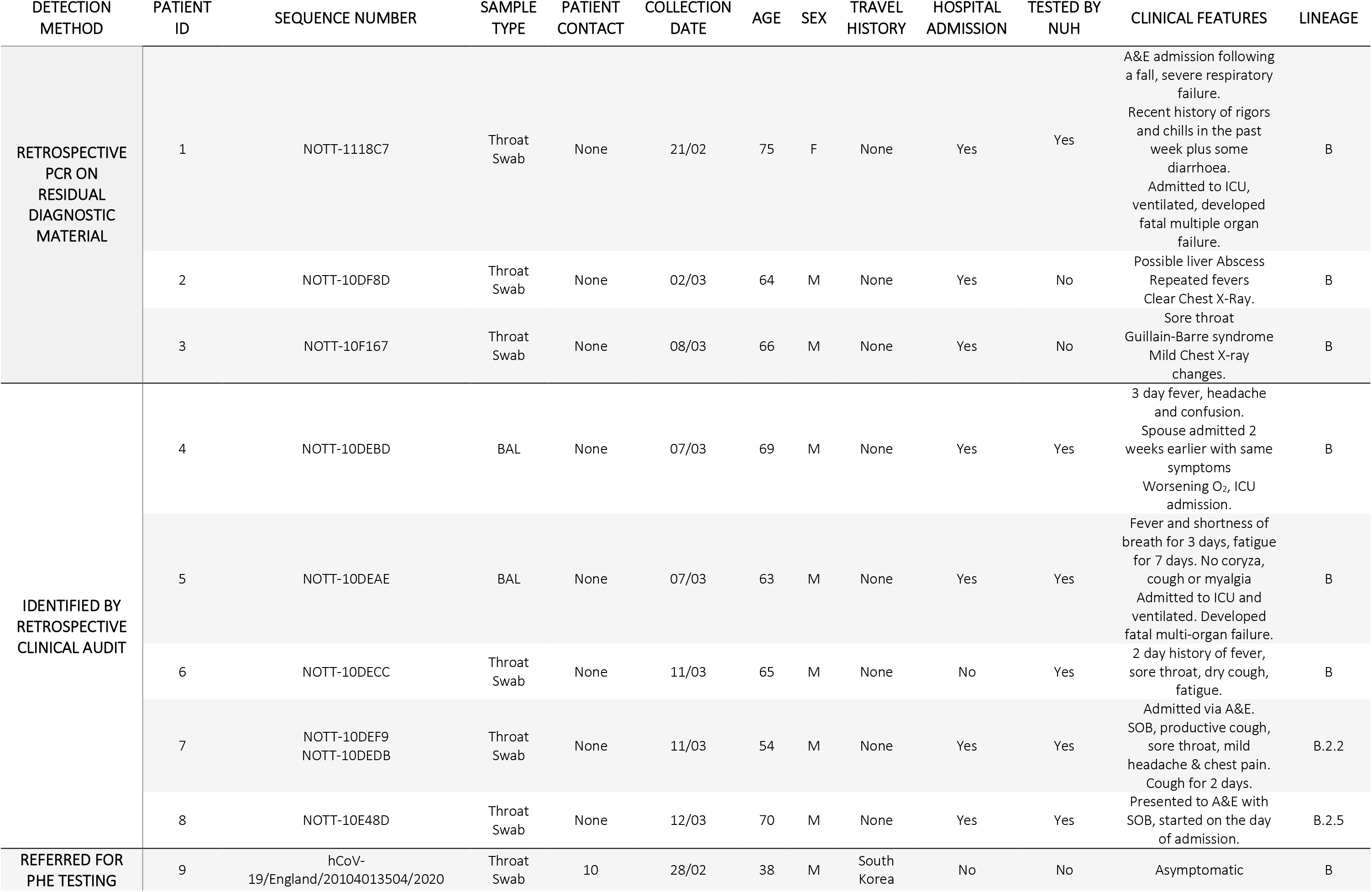
Clinical details for patients in whom SARS-CoV-2 was detected prior to the 12th of March 2020. A&E: Accident & emergency department, ICU: Intensive care unit, SOB: Shortness of breath

A review of case histories revealed an additional five individuals whose symptoms were compatible with COVID-19 but who were not tested as they did not meet the epidemiological criteria of the contemporary case definition (patients 4-8). As these were suspected to have SARS-CoV-2 infection, their respiratory samples were included in the verification of a local SARS-CoV-2 PCR test within the Nottingham diagnostic virology laboratory, and subsequently referred to PHE for confirmatory testing. These patients were all males over the age of 50 whose samples were collected between the 2^nd^ and 12^th^ of March. Complete virus genome sequences were obtained from all 5 patients: three were classified as lineage B (Patients 4-6), one as lineage B.2.2 (Patient 7) and one as lineage B.2.5 (Patient 8).

### Retrospective detection of SARS-CoV-2 in residual diagnostic material

Residual diagnostic material collected prior to the 11^th^ of March, and not subjected to the SARS-CoV-2 testing described above, was retrospectively screened for SARS-CoV-2. The majority of these were throat swabs or nasopharyngeal aspirates, with only a small proportion of nasal swabs, sputum or other samples (Supplementary Figure 1A). The patient demographics indicated a slightly higher inclusion of males than females (Supplementary Figure 1B). The proportion of children under the age of ten were also higher than other ages (Supplementary Figure 1C). All samples had originally been sent to the diagnostic virology laboratory for respiratory virus testing. Overall, 41% of the samples contained at least one respiratory virus, with rhinovirus being the most frequently detected, followed by respiratory syncytial virus and human metapneumovirus (Supplementary Figure 1D). Non-SARS-CoV-2 coronaviruses were detected in 4.16% of samples. An average of 170 samples per week were collected throughout January, February and the first week of March, peaking at 234 samples collected between the 2^nd^ and 8^th^ of March (Supplementary Figure 1E).

Of the 1660 samples tested, three SARS-CoV-2 positive samples were identified through retrospective PCR screening, each from a separate pool. These positive samples were collected from patients on the 21^st^ of February (Patient 1), the 2^nd^ of March (Patient 2) and the 8^th^ of March (Patient 3). All three samples were throat swabs and there were no detectable co-infections with other respiratory viruses. Patient 1 is the earliest known detection of SARS-CoV-2 in Nottingham, collected one week before the previous earliest known positive meeting the contemporary case criteria (Table 1). This patient was a 75-year-old female, admitted to hospital following a fall and suffering from respiratory failure. Her condition worsened, requiring ventilation and the patient ultimately developed multi-organ failure and died on the 3^rd^ of March. No recent travel history or contact with a recently returned traveller was identified during care such that the PHE-defined case definition for SARS-CoV-2 testing at the time was not met. Patient 2 was a 64-year-old male being treated for a suspected liver abscess, presenting with repeated fevers, crackles were noted on lung auscultation but there were no abnormalities on chest X-ray. Patient 3 was a 66-year-old male admitted to hospital with a sore throat and symptoms of Guillain-Barre syndrome, with mild abnormalities also noted on a chest X-ray. Both patients 2 and 3 recovered from their infections.

The complete SARS-CoV-2 genome was sequenced from all 3 samples; all were classified as lineage B.

### Prevalence of SARS-CoV-2 lineages in Nottingham UK following the rollout of localised testing

A total of 1405 patients tested positive for SARS-CoV-2 between the 12^th^ of March and 2^nd^ of June. The total number of positives detected per day peaked on the 8^th^ of April before gradually declining from the 22nd of April onwards (Figure 1).

**Figure 1:**
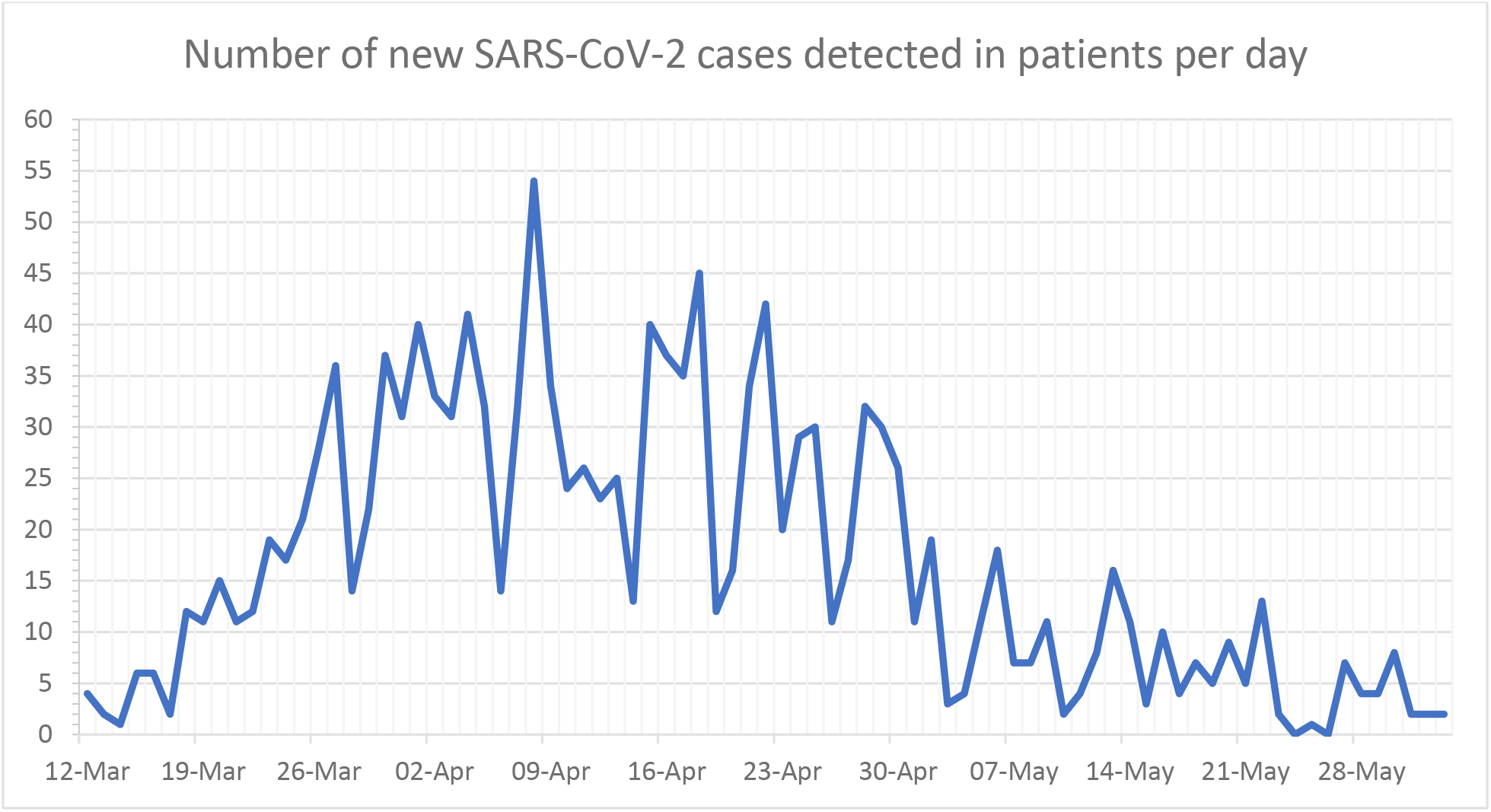
Number of new SARS-CoV-2 Cases Detected in NUH patients per day, from the rollout of localised testing on the 12th of March until the 2nd of June

SARS-CoV-2 positive samples were randomly selected for whole-genome sequencing: of these, 679 were successfully sequenced and classified into lineages. A total of 23 lineages were detected, with lineages B, B.1, B.1.1, B.1.1.1 and B.1.p11 the most frequent (Figure 2A). Lineages B.1, B.11 and B are also the three most prevalent lineages globally (Supplementary figure 2). Between the 2^nd^ of March and 3^rd^ of June 2020, lineages B, B.1.1.1, B.1.1.9, B.1.8, B.1.p11, B.15, B.1.34, B.2.2, B.2.6 and B.6 were proportionally more prevalent in Nottingham than the rest of the UK. Additionally, over 20% of lineage B.1.1.9, B.1.8, B.1.34 and B.6 UK sequences were generated from specimens submitted to Nottingham diagnostic laboratories for testing (Figure 2B). The diversity of lineages detected each week varied over time. Diversity was greatest between week 12 and 15 (16^th^ March to 12^th^ April) and peaked in week 13, during which 19 lineages were detected. The frequency of lineages B.1.1 and B.1.1.1 increased over time, while the other common lineages (B.1 and B.1.p11) decreased in frequency. The three lineages detected prior to the rollout of localised screening - B, B.2.2 and B.2.5 - all decreased in frequency; B.2.2 was last detected in week 17, B.2.5 in week 13 and B in week 21 (Figure 3). Fourteen lineages had not been detected in the month prior to the 2^nd^ of June, 6 of which had not been detected elsewhere in the UK for over a month. Only 5 lineages were detected in the final week of this study (Figure 4).

**Figure 2:**
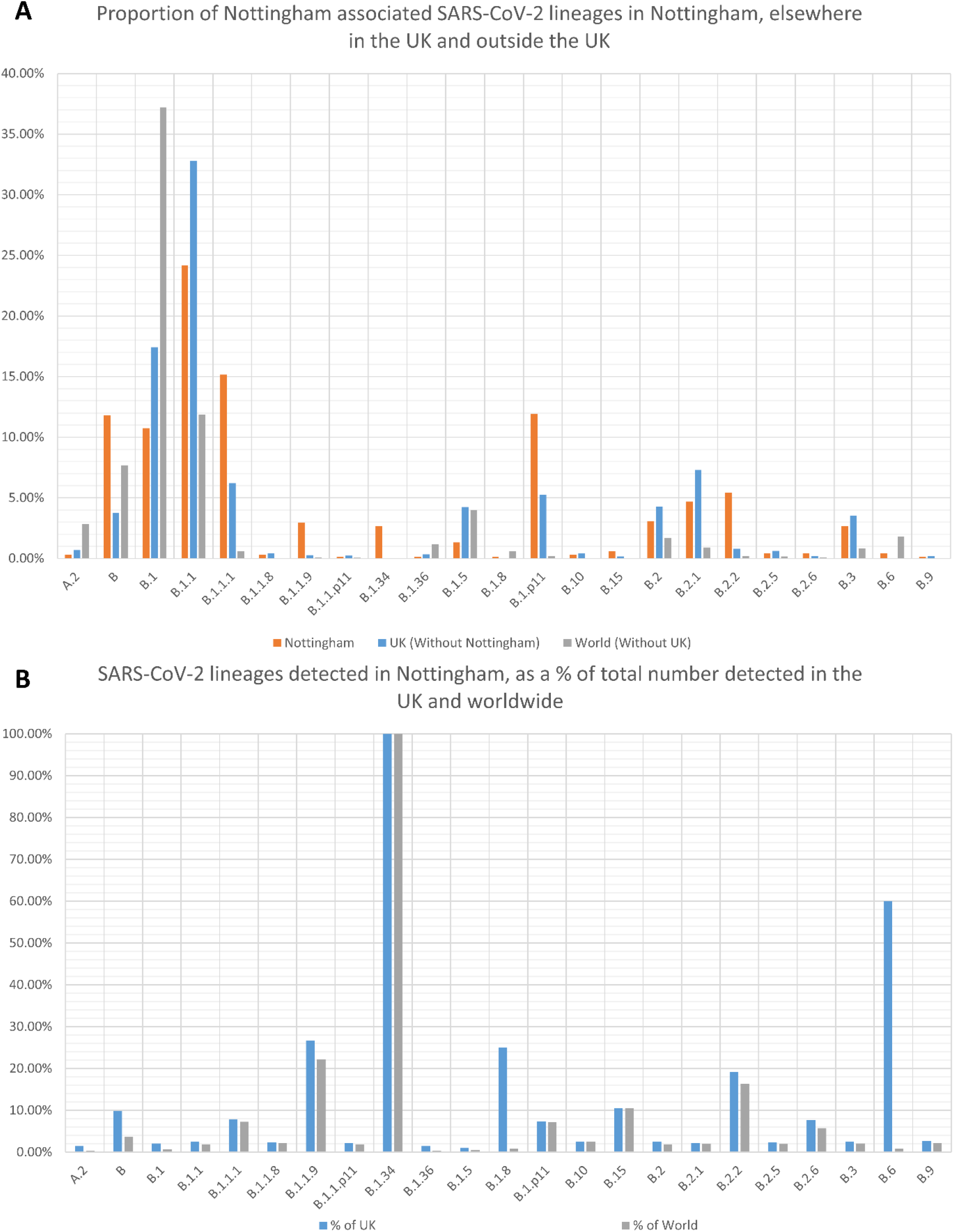
Proportion of SARS-CoV-2 lineages detected between the 21^st^ of February and 2nd of June 2020 in Nottingham, compared with the prevalence of the same lineages in the rest of the UK and elsewhere in the world during the same time period (A). The total count of these lineages is also shown as a percentage of the total count of the same lineages detected in the rest of the UK and world (B).

**Figure 3:**
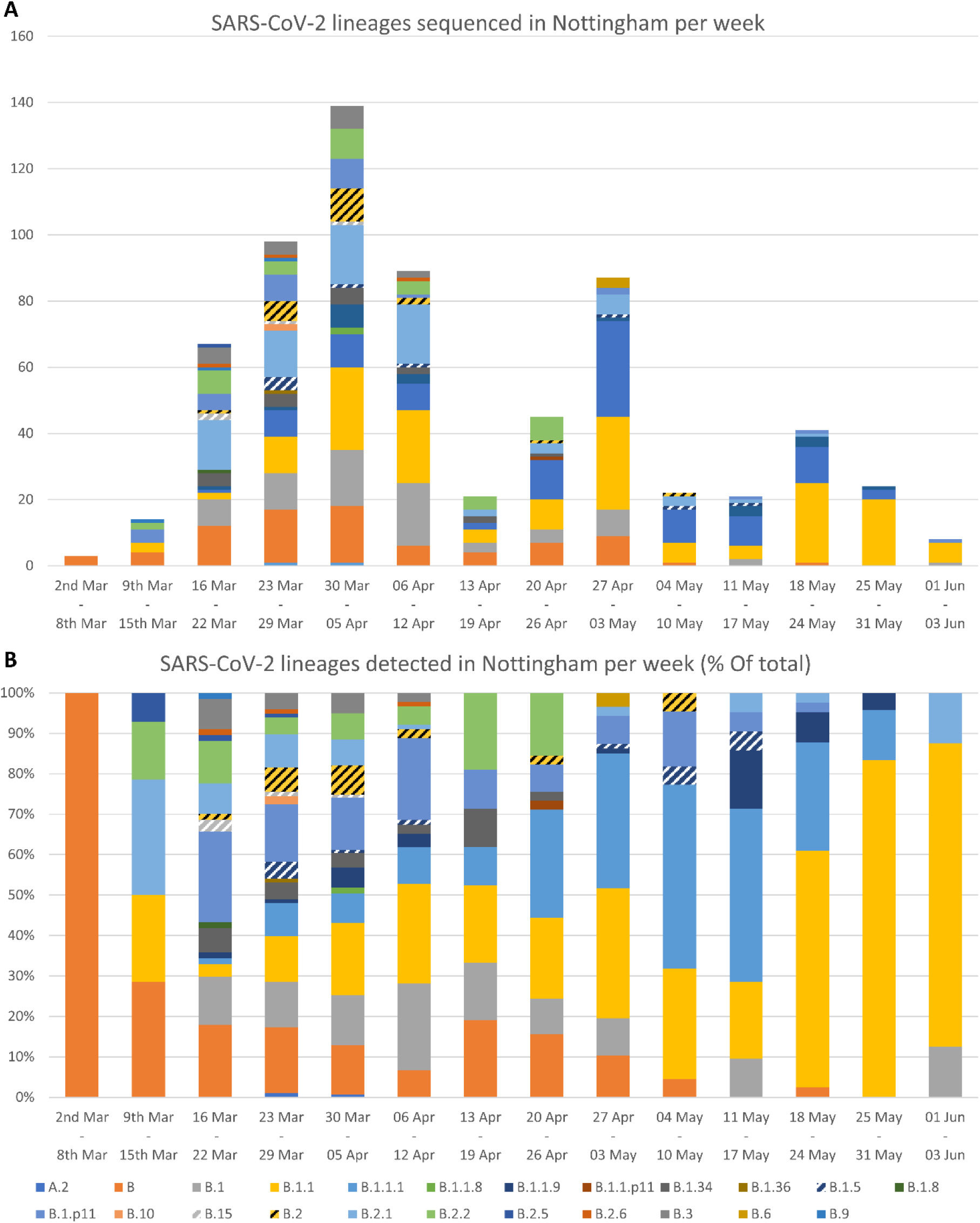
Frequency of SARS-CoV-2 lineages detected in Nottingham between the 21^st^ of February and 2^nd^ of June 2020, per week, as total count (A) and as a proportion of the total number (B).

**Figure 4:**
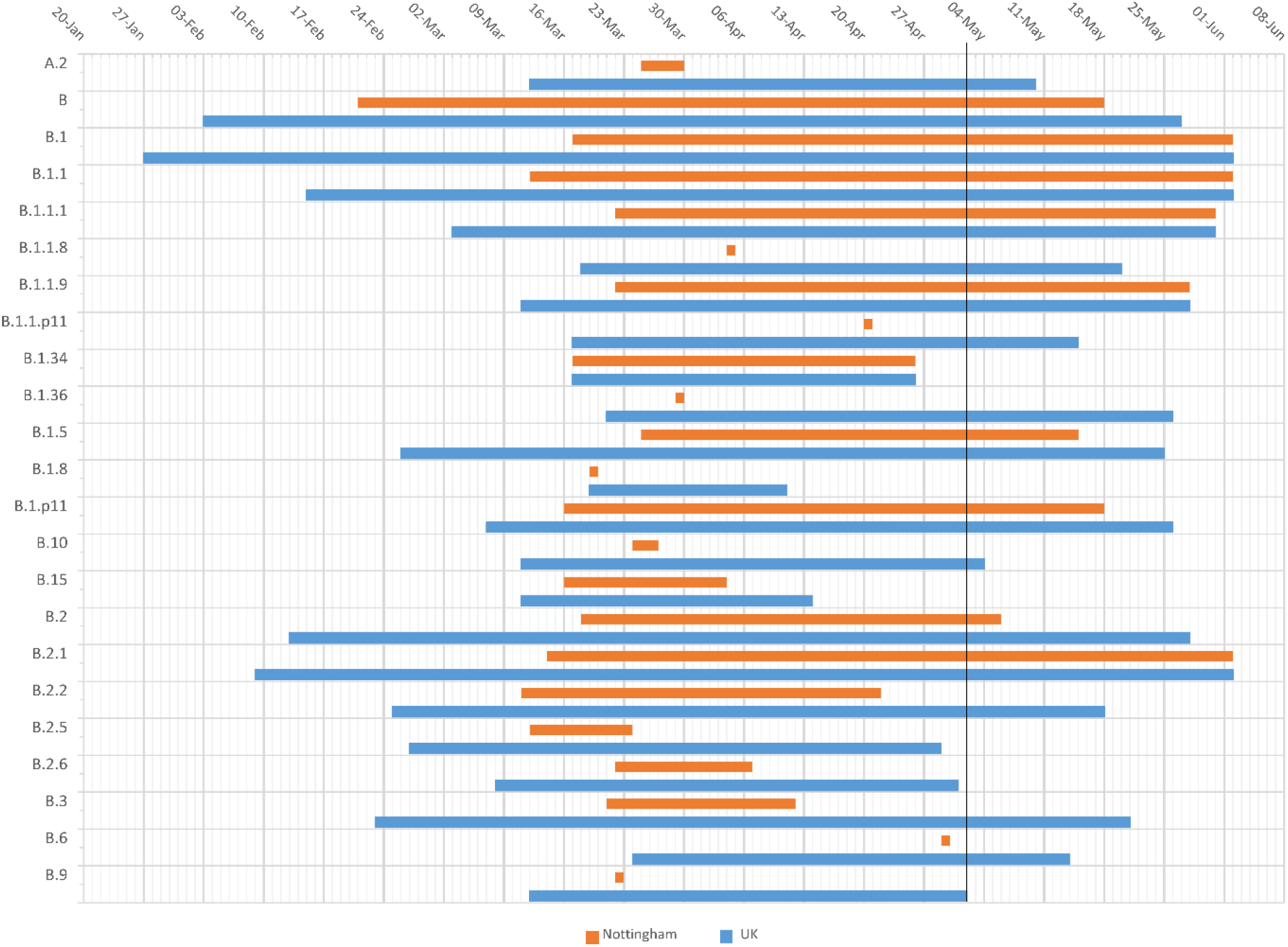
The date in which each lineage was first and last detected in both Nottingham and the UK. The black line marks the 2^nd^ of May, one month prior to the latest available data at the time of writing (2^nd^ of June). Lineages which were last detected before this date are considered to be ‘Unobserved’ ^24^

### Genomic epidemiology of SARS-CoV-2 in Nottingham

Sequences obtained from patients during the retrospective ‘look-back’ analysis of archival respiratory samples were assigned to lineage B using the Pangolin tool. These sequences, together with other lineage B sequences sampled in Nottingham (following the introduction of localised screening), the UK and worldwide were then combined and subjected to phylogenetic analysis.

The resulting phylogeny shows that most lineage B Nottingham-derived sequences form a distinct clade within the B-lineage with strong statistical support (Figure 5, Supplementary figure 3). Sequences collected from other regions in the UK (Sheffield, Cambridge & Exeter) and internationally (Iceland, Portugal, USA) were also interspersed throughout this clade, suggesting an epidemiological connection to those infections in Nottingham. The sequence of patient 1, which was derived from a specimen collected on 21^st^ February, indicates that this individual was part of a transmission chain following the early introduction of this virus strain into the region. Importantly, this sequence clustered with, and predates what was previously thought to be the first case of SARS-CoV-2 infection, which was identified in a traveller returning from South Korea (Patient 8, Table 1).

**Figure 5:**
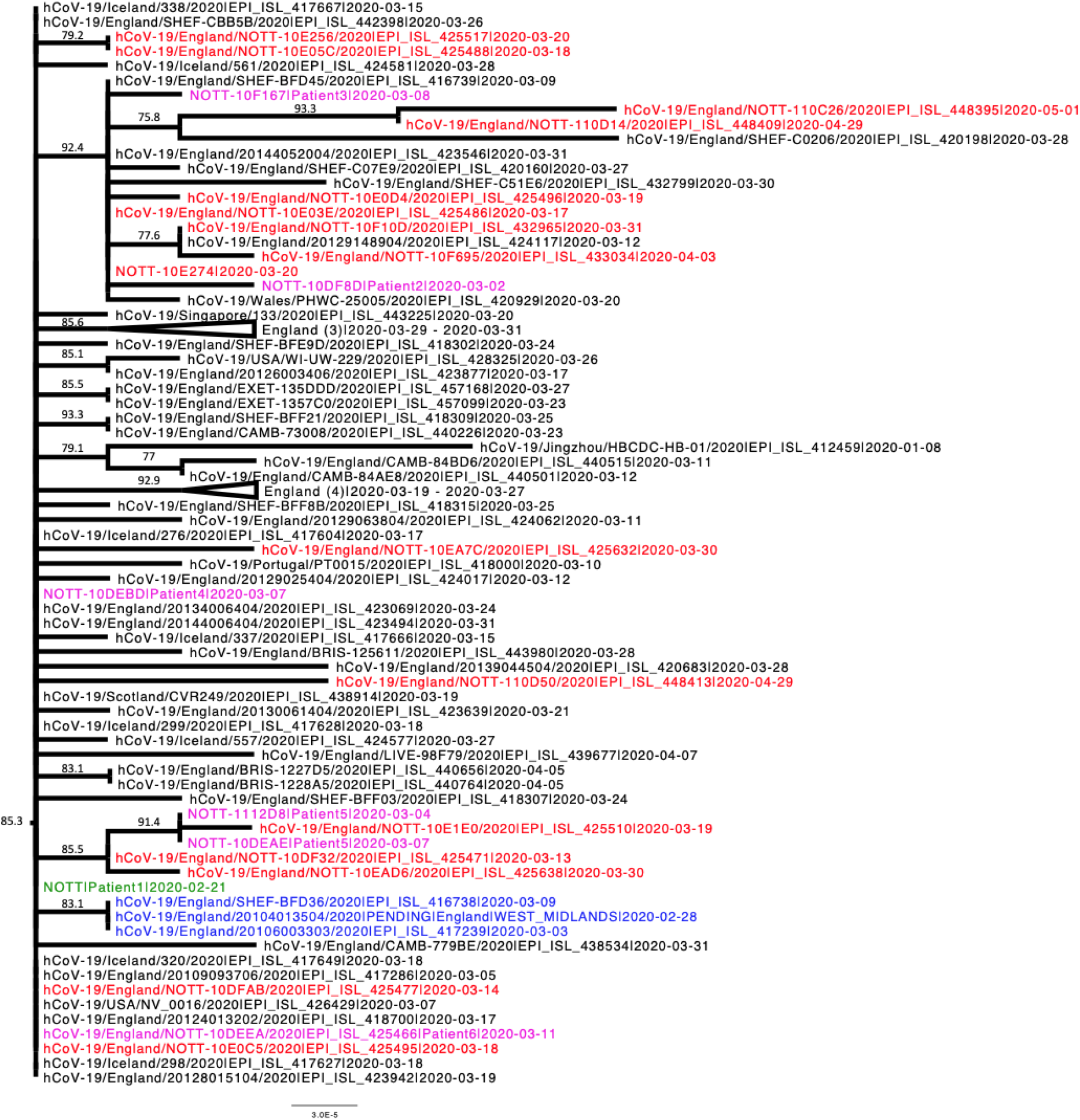
Phylogenetic relationships of SARS-CoV-2 sequences from lineage B based on their entire genome (29,412nt). This is an extraction of a sub-tree that contains most of the lineage B sequences detected in Nottingham, including those identified during retrospective PCR testing. The earliest known sequence is shown in green, other sequences identified in patients during the retrospective analysis are shown in purple, sequences obtained from the first confirmed local case (a traveller returning from South Korea) are shown in blue and sequences obtained from other local patients identified after introduction of wide-screen testing are shown in red. For clarity some branches have been collapsed. Where a branch has been collapsed the geographical locations of where the sequences were obtained are shown at the tip, and the number of individual sequences represented is shown in parentheses. The tree was rooted on a Wuhan sequence sampled on 2020-01-05. Reference sequences are indicated by their GISAID accession numbers. Branch lengths are drawn to a scale of nucleotide substitutions per site. Numbers above individual branches indicate SH-aLRT bootstrap support. The complete, unedited lineage B phylogenetic tree is presented in supplementary figure 3.

Phylogenetic analysis of the remaining Nottingham-derived sequences was performed to determine the number of potential introductions of virus into the Nottingham area in the initial stages of the outbreak. Lineages B.1.5, B.1.36, B.2.6, B.6, B.9 and B.15 were represented by a single sequence, each indicating a unique regional introduction of virus (Supplementary figures 4A-F). Several Nottingham-derived sequences clustered together with sequences representing lineages B.1.34 and B.2.2, again most likely following a single introduction (Supplementary figures 4B & 4G). Analysis of lineages B.1.1.1, B.1.P11, B. 2.5 and B.3 shows that each lineage was independently introduced into Nottingham on at least two occasions (Supplementary figures 4H-K), whilst lineages B.1.1, B.2 and B.2.1 appear to have been introduced at least three times (Supplementary figures 4L-N) and lineage B.1 appears to have been introduced on several occasions (Supplementary figure 4O). Poor branch support in several of these phylogenies, lineage B.1 in particular, precludes the accurate determination of number of introductions, and possibly represents an underestimate of the number of unique introductions. On many occasions, Nottingham sequences clustered with sequences from other UK cities and/or other countries, indicating transmission of the virus via travelling, as well as the potential origins and/or destination of the transmission chains. Interestingly, in lineages B, B.1.1, B.1.1.1, B.1.p11, B.1.34, B.2 and B.2.2, small clusters of Nottingham sequences with high SH-aLRT support were observed (data not shown), suggestive of local community transmission chains of the virus.

## Discussion

The complete genome sequencing of SARS-CoV-2 combined with retrospective clinical and molecular evaluation has facilitated analysis of the introduction and subsequent prevalence of multiple lineages of the virus into Nottinghamshire in the East Midlands region of the UK. Our study reveals multiple community-acquired cases of SARS-CoV-2 lineage B viruses presenting at a regional healthcare centre, but failing to meet contemporary case criteria for testing in a diagnostic system with restricted capability and up to one month before the government imposed countrywide lockdown measures.

Patient 1 in this study is, to the best of our knowledge, the earliest described community-acquired case of SARS-CoV-2 in the UK, admitted to hospital care on the 21^st^ of February 2020, and was also the first UK COVID-19 death, preceding the earliest known death by 2 days ^25^. A median incubation period estimate of COVID-19 of 5.1 days (4.5-5.8) from infection to the presentation of initial symptoms ^26^, in addition to the week prior to hospitalisation when the patient was also symptomatic, suggests that infection could have taken place as early as the 9^th^ of February. This patient had no history of either travel or contact with travellers and so infection must therefore have occurred locally, suggesting an active wider network of community transmission than previously suspected. The first PHE-confirmed case of SARS-CoV-2 in Nottinghamshire meeting contemporary clinical testing criteria, patient 9, was sampled one-week after patient 1. This patient had recently travelled to South Korea, but their associated viral sequence belonged to the same clade of lineage B as patient 1, which has been infrequently observed in global data sets. The patient developed a fever 5 days after returning to Nottingham, which is consistent with both acquisition in South Korea and local acquisition in the UK before or after international travel ^26^. Locally acquired infection in Nottingham is probably the most likely scenario, and certainly the most parsimonious explanation, given our evidence that this viral lineage was already circulating in the Nottingham area. This highlights the utility of next-generation sequencing and phylogenetic analysis in delineating the epidemiology of virus outbreaks.

Whilst initially common, the proportion of lineage B in Nottinghamshire fell sharply as the pandemic continued. At the time of writing, lineage B was last detected on the 18^th^ of May in Nottingham and using the same nomenclature proposal in which lineages were defined, lineages are classed as “unobserved” after a month of not being detected ^24^.

The frequency of SARS-CoV-2 detection and lineage diversity in Nottingham began to increase during mid-March and peaked during late March/early April. This is consistent with the findings of Pybus *et al* ^15^, where the frequency of TMRCAs (Time of the most recent common ancestor), and therefore transmissions, in the UK peaks during late March. Some virus lineages appeared to have been introduced on multiple occasions and were transmitted widely, whereas others had fewer introductions and/or were associated with limited onward spread. Despite the unprecedented national and global effort, only a fraction of SARS-CoV-2 infected individuals were diagnosed and sequenced, with further loss of sequence information due to a high-quality threshold, such that many novel introductions are likely to be undetected. The complete genome sequence data sets from which the phylogenetic analyses were derived are heavily biased towards UK samples, due to unevenness in global sequencing activity: 53% of available genomes were sampled in the UK ^27^, despite accounting for only 3.4% of reported cases^2^. This imbalance will hinder inferences about the definitive sources of introduction to the UK, but conversely has allowed greater insight into subsequent national SARS-CoV-2 transmission within the UK and onwards to other destinations.

It is likely that the true prevalence of the virus within the local study setting and indeed the wider UK during the timeframe of this study was much higher than we have reported here, particularly from early-February onwards, as our study was limited to symptomatic individuals requiring secondary medical care. Asymptomatically infected individuals, along with those presenting with mild or paucisymptomatic infection, are likely to comprise a significant proportion of the total number of infections. Prevalence of asymptomatic infections in representative American and Icelandic populations is high: between 40% and 45% of total infections ^28^, and as high as 87.5% in younger cohorts ^29^. Epidemiological modelling has predicted that for every hospitalised COVID-19 case in the UK, there was a further 120-124 infected individuals ^30^, supporting the hypothesis of a significant undetected rate of infection within the wider community.

Although a large cohort of surplus diagnostic material dating back to January 2020 was screened, it is possible that further SARS-CoV-2 positives within remain undetected. The pooling of respiratory samples, for example, may have contributed to a decrease in the detection rate. However, we consider this unlikely as similar pooling of samples has been shown to result in only a minor increase in reported Ct-values ^31^. Also, the retrospective screening of surplus diagnostic material only captures respiratory samples taken from a specific subgroup of patients: those admitted to hospital predominantly with a suspected respiratory infection.

The salient finding from our study is that simple opportunities to identify early cases of SARS-CoV-2 infection were missed due to overly stringent case-criteria. Had the diagnostic criteria been widened earlier to include patients with compatible symptoms but no travel history, it is likely that earlier imported infections would have been detected, enabling rapid deployment of infection control measures that may have prevented onwards transmission. However, the diagnostic capacity available nationally was not sufficient at the time to process the volume of testing required with a broader case definition. This would have been ameliorated by increased local testing within PHE and NHS diagnostic laboratories earlier in the epidemic. Many of these laboratories possessed the knowledge and experience necessary to initiate local testing but were unable to do so due to a delayed mandate and the availability of commercial testing platforms, on which many diagnostic services rely. For future pandemic preparedness, the UK urgently needs to invest in and expand diagnostic capacity within NHS and PHE diagnostic laboratory services, ensuring the rapid dissemination of diagnostic protocols, a stable supply chain of reagents and instruments required to sustain a rapid increase in capacity and provide a fully integrated end-to-end result system to ensure appropriate and timely infection prevention and control measures can be taken within secondary care and community settings. Any lasting investment in the human resources and associated infrastructure to achieve a more agile epidemic response both nationally and globally will undoubtedly save lives and drastically reduce the adverse impact of such outbreaks on the economy.

## Data Availability

All sequence data used in this study is available through the COG-UK consortium and GIS-AID initiative.

## Author Contributions

The study was designed by C.P.M, J.G.C and J.K.B. Clinical specimens and details were provided by G.C, M.K, V.M.F, M.M.L, H.H-W, W.S, T.C.J.B, L.B, I.W, A.J and W.L.I. Retrospective RT-PCR screening and preparation of samples for sequencing was completed by J.G.C and C.P.M. Complete genome sequencing was performed by M.L, N.H, C.M, M.C, F.S, J.D, and V.M. Data was analysed by T.T, E.H, C. P.M, J.G.C, M.L and J.K.B. The manuscript was written by J.G.C and revised by C.P.M, J.K.B, T.T, W.L.I, L.B, G.C, E.H and M.L. All authors read the final manuscript.

## Acknowledgements

This work was supported by grants from the Medical Research Council UK (MR/R010307/1 and MR/S009434/1), which are both part of the EDCTP2 programme supported by the European Union, The University of Nottingham Campaign and Alumni Relations Office research donations award and the EU H2020 programme (Project 727393 - PaleBlu). Whole genome sequencing of SARS-CoV-2 was funded by COG-UK; COG-UK is supported by funding from the Medical Research Council (MRC) part of UK Research & Innovation (UKRI), the National Institute of Health Research (NIHR) and Genome Research Limited, operating as the Wellcome Sanger Institute. We would also like to thank Prof Oliver Pybus, University of Oxford for helpful discussion.

## References

1 Lu H, Stratton CW, Tang YW. Outbreak of pneumonia of unknown etiology in Wuhan, China: The mystery and the miracle. J Med Virol 2020; 92: 401–2.

2 WHO Coronavirus Disease (COVID-19) Dashboard. https://covid19.who.int/ (accessed May 10, 2020).

3 Chen N, Zhou M, Dong X, et al. Epidemiological and clinical characteristics of 99 cases of 2019 novel coronavirus pneumonia in Wuhan, China: a descriptive study. Lancet 2020; 395: 507–13.

4 Huang C, Wang Y, Li X, et al. Clinical features of patients infected with 2019 novel coronavirus in Wuhan, China. Lancet 2020; 395: 497–506.

5 Matthay MA, Aldrich JM, Gotts JE. Treatment for severe acute respiratory distress syndrome from COVID-19. Lancet Respir. Med. 2020; 8: 433–4.

6 Treibel TA, Manisty C, Burton M, et al. COVID-19: PCR screening of asymptomatic health-care workers at London hospital. Lancet. 2020; 395: 1608–10.

7 Rivett L, Sridhar S, Sparkes D, et al. Screening of healthcare workers for SARS-CoV-2 highlights the role of asymptomatic carriage in COVID-19 transmission. Elife 2020; 9. DOI:10.7554/eLife.58728.

8 Rothe C, Schunk M, Sothmann P, et al. Transmission of 2019-NCOV infection from an asymptomatic contact in Germany. N. Engl. J. Med. 2020; 382: 970–1.

9 Wei WE, Li Z, Chiew CJ, Yong SE, Toh MP, Lee VJ. Presymptomatic Transmission of SARS-CoV-2 - Singapore, January 23-March 16, 2020. MMWR Morb Mortal Wkly Rep 2020; 69: 411–5.

10 Bai Y, Yao L, Wei T, et al. Presumed Asymptomatic Carrier Transmission of COVID-19. JAMA - J. Am. Med. Assoc. 2020; 323: 1406–7.

11 Wölfel R, Corman VM, Guggemos W, et al. Virological assessment of hospitalized patients with COVID-2019. Nature 2020;: 1–5.

12 Arons MM, Hatfield KM, Reddy SC, et al. Presymptomatic SARS-CoV-2 Infections and Transmission in a Skilled Nursing Facility. N Engl J Med 2020; published online April 24. DOI:10.1056/nejmoa2008457.

13 Lillie PJ, Samson A, Li A, et al. Novel coronavirus disease (Covid-19): The first two patients in the UK with person to person transmission. J. Infect. 2020; 80: 578–606.

14 Spiteri G, Fielding J, Diercke M, et al. First cases of coronavirus disease 2019 (COVID-19) in the WHO European Region, 24 January to 21 February 2020. Eurosurveillance. 2020; 25. DOI:10.2807/1560-7917.ES.2020.25.9.2000178.

15 Pybus O, Rambaut A, Zarebski AE, et al. Preliminary analysis of SARS-CoV-2 importation & establishment of UK transmission lineages 8. 2020.

16 GOV.UK. COVID-19: investigation and initial clinical management of possible cases. Gov.Uk. 2020;: 1-5.

17 Tsang TK, Wu P, Lin Y, Lau EHY, Leung GM, Cowling BJ. Effect of changing case definitions for COVID-19 on the epidemic curve and transmission parameters in mainland China: a modelling study. Lancet Public Heal 2020; 5: e289–96.

18 An integrated national scale SARS-CoV-2 genomic surveillance network. The Lancet Microbe 2020; 0. DOI:10.1016/s2666-5247(20)30054-9.

19 Bagasi AA, Howson-Wells HC, Clark G, et al. Human Bocavirus infection and respiratory tract disease identified in a UK patient cohort. J Clin Virol 2020;: 104453.

20 GitHub - artic-network/artic-ncov2019: ARTIC nanopore protocol for nCoV2019 novel coronavirus. https://github.com/artic-network/artic-ncov2019 (accessed May 27, 2020).

21 O’Toole A, McCrone J. hCoV-2019/pangolin: Software package for assigning SARS-CoV-2 genome sequences to global lineages. 2020. https://github.com/hCoV-2019/pangolin (accessed June 21, 2020).

22 Minh BQ, Schmidt HA, Chernomor O, et al. IQ-TREE 2: New Models and Efficient Methods for Phylogenetic Inference in the Genomic Era. Mol Biol Evol 2020; 37: 1530–4.

23 Guindon S, Dufayard JF, Lefort V, Anisimova M, Hordijk W, Gascuel O. New algorithms and methods to estimate maximum-likelihood phylogenies: Assessing the performance of PhyML 3.0. Syst Biol 2010; 59: 307–21.

24 Rambaut A, Holmes EC, Hill V, et al. A dynamic nomenclature proposal for SARS-CoV-2 to assist genomic epidemiology. *bioRxiv* 2020;: 2020.04.17.046086.

25 CMO for England announces first death of patient with COVID-19 - GOV.UK. https://www.gov.uk/government/news/cmo-for-england-announces-first-death-of-patient-with-covid-19 (accessed Aug 5, 2020).

26 Lauer SA, Grantz KH, Bi Q, et al. The incubation period of coronavirus disease 2019 (CoVID-19) from publicly reported confirmed cases: Estimation and application. Ann Intern Med 2020; 172: 577–82.

27 Argimón S, Abudahab K, Goater RJE, et al. Microreact: visualizing and sharing data for genomic epidemiology and phylogeography. Microb genomics 2016; 2: e000093.

28 Oran DP, Topol EJ. Prevalence of Asymptomatic SARS-CoV-2 Infection: A Narrative Review. Ann Intern Med 2020; published online June 3. DOI:10.7326/M20-3012.

29 Lytras T, Dellis G, Flountzi A, et al. High prevalence of SARS-CoV-2 infection in repatriation flights to Greece from three European countries. J Travel Med 2020. DOI:10.1093/jtm/taaa054.

30 Jit M, Jombart T, Nightingale ES, et al. Estimating number of cases and spread of coronavirus disease (COVID-19) using critical care admissions, United Kingdom, February to March 2020. Euro Surveill 2020; 25: 4–8.

31 Lohse S, Pfuhl T, Berkó-Göttel B, et al. Pooling of samples for testing for SARS-CoV-2 in asymptomatic people. Lancet Infect Dis 2020; 3099: 2019–20.

